# Expanding Faculty Representation in US Academic Neurological Surgery: Achievements and On-going Challenges

**DOI:** 10.64898/2026.04.24.26351672

**Authors:** Jack M. Shireman, Neel Mukherjee, Krista Brackman, Nathan Kurtz, Akshita Patniak, Liam McCarthy, Nikita Gonugunta, Simon Ammanuel, Mahua Dey

## Abstract

**Objectives:** Academic medical institutions are the gatekeepers of the physician workforce and shape the future of medicine by regulating medical school admissions as well as residency training. Although broadly the field of medicine is seeing more representation from traditionally underrepresented groups, the critical decision-making platform of academic medicine continues to be uncharacteristically homogeneous, represented mainly by white males. This is even more pronounced in surgical subspecialties, such as academic neurosurgery. This study aims to quantify this phenomenon, uncover its driving factors, and define opportunities for improvement.

**Methods:** Using a mixed research methodology, academic neurosurgical faculty in the U.S were identified, and their demographic data was collected. An internet search using Google Scholar and Scopus was conducted to determine scholarly activity using number of publications and h-index.

**Results:** We found a significant increase in female faculty in academic neurosurgery within the last decade. Comparing the faculty rank amongst male and female faculty, we found that the majority of female faculty are at the assistant professor level (n=36/79; 45.6%) while male faculty are more at the full professor rank (n=265/582; 45.5%). A similar trend was seen for under-represented minority neurosurgery faculty. Strong scholarly activity corelated with a departmental chair position for male faculty, however, this trend was not true for female faculty. There was a significant difference in the number of publications and h-index in female vs male faculty, but only when including male faculty outliers at the full professor level.

**Conclusion:** Slowly but steadily, academic neurosurgery is making progress towards a more diverse and representative workforce in the U.S that better reflects the patient population. Facilitating timely progression of females and URM neurosurgeons into senior professorship and academic leadership roles will further advance this essential progress.

## Introduction

Diversity in healthcare has been shown to improve patient outcomes [1–10]. When a patient is involved with a diverse healthcare team, they are more likely to receive an accurate diagnosis, experience higher patient satisfaction, adhere properly to treatment plans, and participate in preventative measures [5,11]. Diverse clinical teams also allow for representation of a broader range of perspectives which improves decision making and can be particularly impactful in managing culturally nuanced care scenarios such as end-of-life care or overcoming language barriers [10]. Many in the medical field have begun to recognize these gaps and have called for a push in initiatives aimed at increasing representation, however, these initiatives have struggled to make meaningful change in surgical specialties which remain largely homogenous across both gender and race [12–15]. Traditionally, surgical specialties presented unique challenges in attracting and retaining diverse talent due to their notoriously time-intensive and rigid residency structures, which can make it harder to navigate activities such as family planning, childbirth, or caregiver responsibilities [16]. This rigid residency structure and slow search/adaptation of innovative changes in educational methodology likely had a disproportionate effect on women who are balancing real-life decisions on childbirth and childcare with the time commitment of a long residency [17]. However, this is a very simplistic explanation of a very complex societal phenomenon. Many systematic regulatory changes have been made in medicine to address demographic disparities over last decades, but their outcomes have been mixed [7,9,12,14,15,18–21].

Although several specialties have benefited from these changes and made remarkable strides towards attaining and sustaining racial and gender balance, there are still several specialties such as neurosurgery that are disproportionately homogenous and lagging general trend [19,22–25] . Recent studies have shown that lack of gender representation worsens with vertical movement from trainee level to the higher-level ranks, such as full professorship to departmental leadership and beyond [20,24]. This trend is especially pertinent in the subsect of academic neurosurgery, where female neurosurgeons comprise only 8-12% of the workforce [19,20].

The reasons for unequal outcomes across specialties from similar regulatory practices are complex and multifactorial. They include structural barriers limiting early access and exposure to certain specialties as well as the human aspect of social networking principles like homophily, the tendency of individuals to associate/work with similar individuals across race or gender. These often under recognized deeply rooted societal factors can manifest in the form of slower development and advancement of minority faculty and result in increased burnout, decreased morale, and high drop-out rates, forming a self-fulfilling negative loop. These factors are often felt more by minority groups, however, are not studied and well documented in the literature in an objective data driven manner.

Academic neurosurgery has always been at the forefront of driving change and innovation. It is also a critical area for studying and understanding diversity and inclusion patterns, as it has a direct impact on the accepted norms for the specialty and dictates all training requirements and competencies that must be met to enter the profession. Over the last two decades, several calls to action have documented slow forward progress in terms of recruiting more female residents in the field, these numbers drop significantly when tracking future development in leadership positions such as academic faculty appointment, departmental leadership, professional society leadership, professional achievement awards [18,23]. Objectively, progress in academia is closely related to scholarly activity, we collected demographic data and scholarly activity (number of publications and H-index) from nearly 700 academic neurosurgery faculty and examined what factors drive academic advancement.

## Methods

### Neurosurgery Residency Program Screening

Using the neurosurgical residency training program directory provided by the American Association of Neurological Surgeons (https://www.aans.org/en/Trainees/Residency-Directory), a list of all institutions in the United States with a neurosurgery residency program was compiled. Neurosurgery faculty names and emails from the institutions’ websites were recorded. For the purposes of this study, neurosurgery faculty was defined as a person who completed neurosurgery residency training and maintains a clinically active, academic neurosurgery position at a medical institution in the United States. Recorded emails were used to disperse the Diversity in Neurosurgery survey which was administered using the Qualtrics survey platform (Supplementary Table 1). Academic rank, gender, and race were obtained from publicly available academic institution websites for faculty who did not respond to the survey. Survey information was tracked but all data reported or released were deidentified and coded to maintain privacy. (Figure 1A)

**Figure 1.**
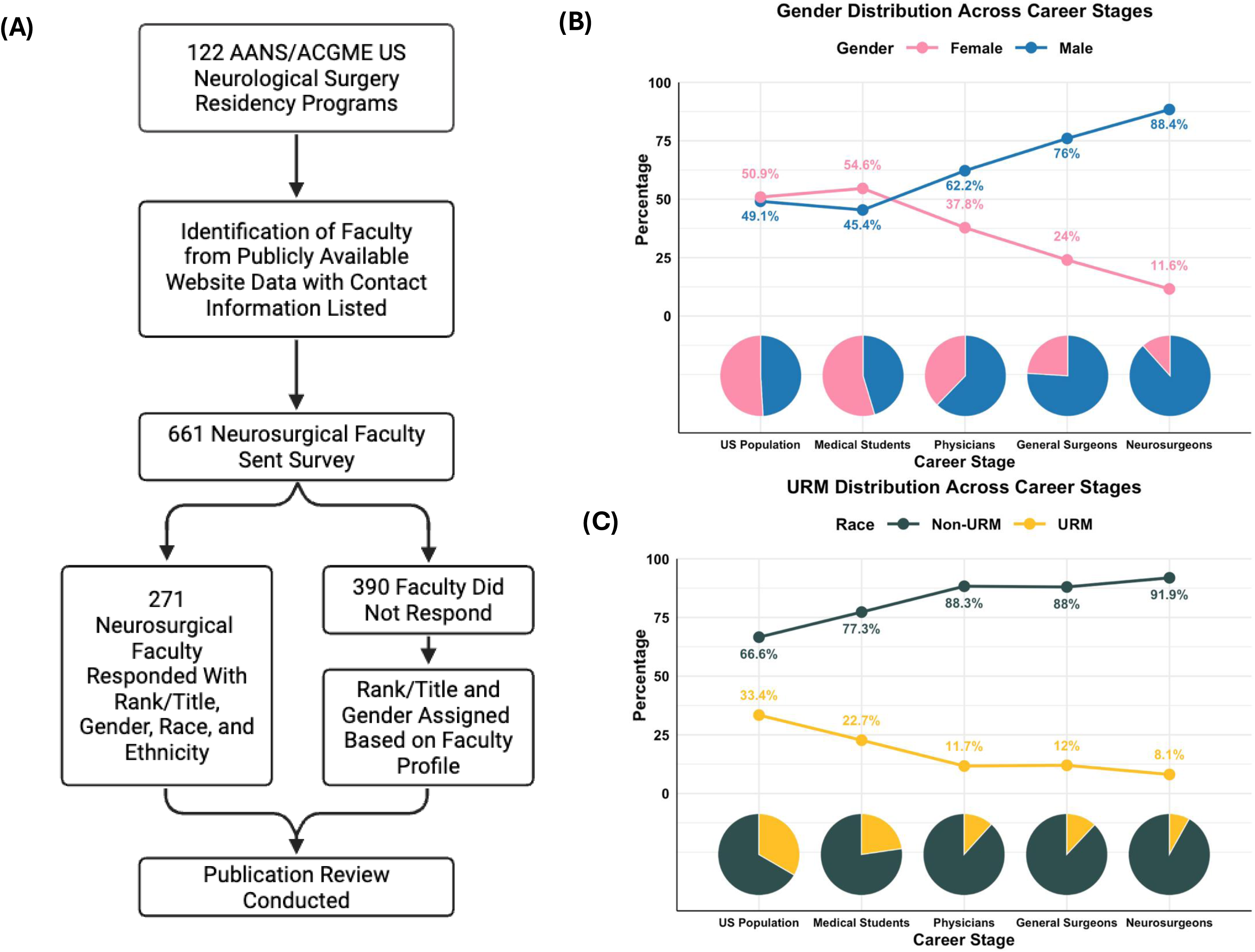
Gender and racial/ethnic disparity throughout the neurosurgery pipeline and overview of survey cohort development: **A)** Flowchart outlining the inclusion and categorization process for neurosurgical faculty based on survey response or publicly available data. **B)** Line and pie chart visualization of gender distribution across five stages of neurosurgical specialization. **C)** Line and pie chart visualization of racial/ethnic distribution across five stages of neurosurgical specialization.

### Internet Scrape

A publication review was conducted on all 661 identified academic neurological faculty regardless of response to the survey using PubMed advanced search. The review only included publications in journals that were indexed by PubMed. Total number of publications was recorded and further analyzed to group the publications into first, last, and middle authorship categories. Preferred names from the survey were also used whenever possible to improve publication identification. H-index was documented via Scopus using the respondent’s name and affiliated institution. At the end of data collection, the results were spot checked by both NM and JMS for accuracy. The publication review and data collection comprise data current up to August of 2023.

For information on the racial demographics, we relied solely on the data from the respondents to the survey because it would be inappropriate to accurately assign race information from publicly available data on faculty webpages. For ease of reporting, we classified race into two categories URM and non-URM (as defined by the AAMC) with URM including African American, Hispanic/Latino, and Native American/Pacific Islander self-reported races.

### Statistical Analysis

All statistics were conducted using either GraphPad Prism or R. For comparison between 2 groups students T-test was conducted while comparison across more than 2 groups utilized an ANOVA + Tukey’s multiple comparison. Statistical significance was considered as a p-value < 0.05 and is reported in the results section and figure legends. All cartoons were created using Biorender.

## Results

### There are significantly larger gender and racial disparities in neurosurgery compared to all physicians and general surgeons

To contextualize the survey data, we utilized US census data and medical school admissions and physician employment data (obtained from the AAMC) to break down the gender and racial balance through stages of academic neurosurgical training (Figure 1B & C). At the US population level gender balance is roughly equal with females holding a slight advantage of roughly 1% (Figure 1A). This balance tips even more heavily at the level of enrolled medical students where females hold roughly 55% of medical school class seats compared to males at 45% (Figure 1A). When examining data on all current physicians, the gender balance flips wildly back towards males (62% vs 38%) and only worsens at the level of general surgeons (81.5% to 18.5%) and neurosurgeon (88% to 12%) (Figure 1B). In terms of race, we classified individuals from our survey and the general population as URM if they are self-reported African American, Native American/ Pacific Islander, or Hispanic/Latino. By this definition the URM population in medical schools, as practicing physicians, as practicing surgeons, and as academic neurosurgeons was even more imbalanced than our gender data (Figure 1C). In fact, in our survey population over 90% of the academic neurosurgeons identified as non-URM (Figure 1C).

### A greater proportion of female and URM faculty are at the assistant professor level compared to non-URM male faculty

Promotion through the ranks of faculty at institutions is generally thought of as a marker for career success and upward trajectory. Critically, these senior full professor appointments often also come with leadership roles within the department and the job security that is granted by faculty tenure. We examined our survey data to understand the gender balance of neurosurgery faculty across the three promotional levels and found that males outnumber females at all levels (Figure 2A). When accounted for in terms of percent of total gender represented, we found that a higher proportion of females occupied assistant professor roles than males (45% female to 29% male), while a higher number of males occupied full professor levels than females (46% male to 33% female) (Figure 2B). For URM faculty, our survey data shows only single digit levels of URM academic neurosurgeons at all promotional levels with most URM faculty at the Assistant Professor level compared to the full professor level (Figure 2B &C). These data demonstrate a slower transition from assistant to full professor for both female and URM faculty.

**Figure 2.**
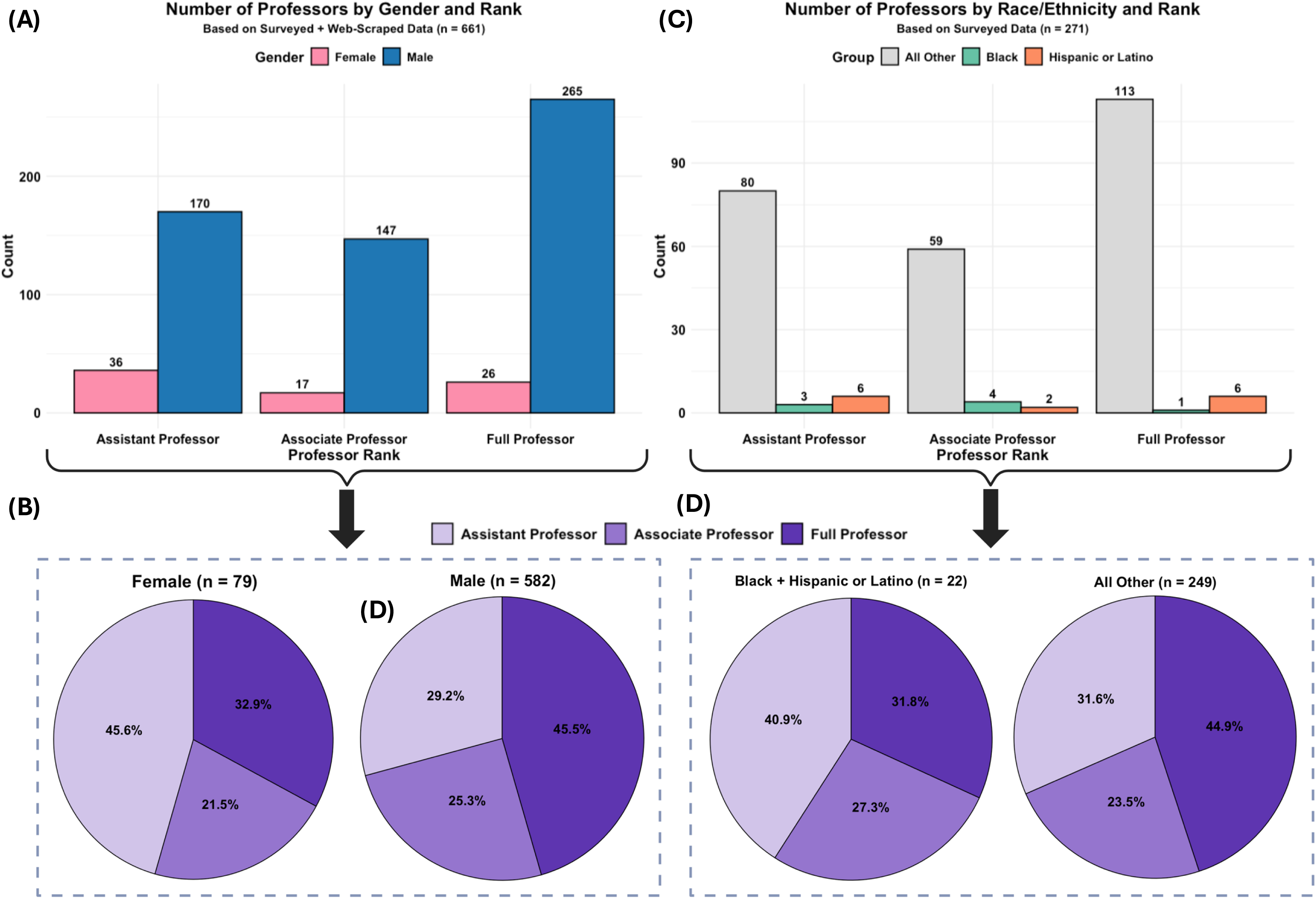
Rank distribution by gender and race among neurosurgery faculty: **A)** Bar plot showing the number of male and female neurosurgeons at each academic rank using combined survey and web-scraped data. **B)** Pie chart showing the proportion of faculty by rank across gender. **C)** Bar plot showing the number of neurosurgeons in each given racial/ethnic group at each academic rank using only surveyed data. **D)** Pie chart showing the proportion of faculty by rank across race.

### Scholarly accomplishments corelate with more leadership positions in male faculty compared to female faculty

A main driving factor for promotion throughout academia is the publication of scientific literature. Examining total publications and h-index across different faculty rank levels revealed a significant difference between genders was present only at the level of full professor for both h-index and total publications (Figure 3A). This change was so large it drove statistical significance when lumping all academic levels together in both the number of publications and h-index between male and female neurosurgical faculty (p < 0.001 at both levels) (Figure 3B). Because of the significant outliers present in this analysis we isolated and compared scholarly activity of the top 15 male and female faculty. We utilized a 70/30 weighing scheme of h-index and total publications to account for not only high publication volume but also impactful and highly cited articles in the field (Figure 3C). Full professor positions in individuals with a high h-index and strong publication record typically indicate a leadership position within the department. We utilized faculty webpages to search for individuals who either are or had been a Chair for their department (Figure 3C, indicated by triangles). This data revealed that most of the male faculty (11/15) with high scholarly activity either are or were chairs of a neurosurgery department while only (3/15) of the female faculty with high scholarly activity by the same metrics had attained similar leadership positions (Figure 3C). After removing the 15 most prolific publishing male faculty from the cohort the statistical significance of the comparison between males and females for total publications and h-index is no longer observed (Figure 3D).

**Figure 3.**
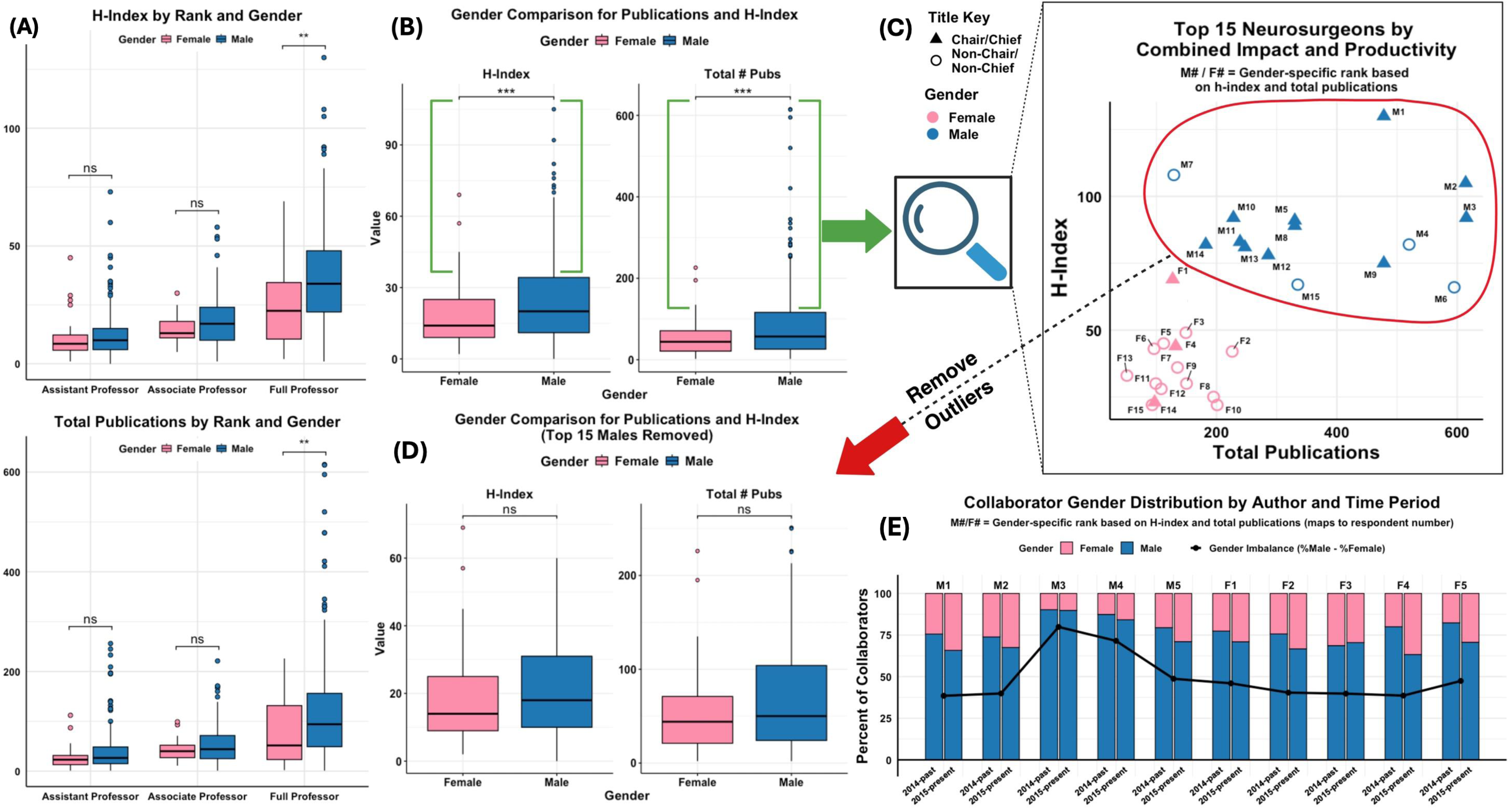
Gender disparities in academic productivity and collaboration among neurosurgeons with emphasis on top performers: **A)** Bar plots depicting h-index (above) and total publications (below) by gender within each academic rank. **B)** Boxplots comparing h-index and total publications by gender. Green brackets highlighting the presence of outliers. **C)** Scatterplot of top 15 male and top 15 female neurosurgeons ranked by combined h-index and publication count. Red outline depicts clustering of males. **D)** Re-analysis of h-index and total publications after removal of top 15 males. **E)** Stacked bar plots showing the gender composition of co-authors for the top five male and female neurosurgeons across two time periods. Black line tracks gender imbalance in collaboration.

We hypothesized that one of the reasons for the gap in transition to full professor, and therefore department leadership roles, among females was the phenomenon of homophily, that male faculty already at the top of the field were more likely to collaborate with other male faculty. To test this, we examined publications from the top 5 male faculty and female faculty in terms of our weighted academic productivity metric from figure 3C in the decades of 2014-past and 2015-present. We utilized the open-source gender-guesser python package to assign a male vs female collaboration score. We found an overall bias towards male collaborators across both male and female professors (Figure 3E). We also found that although females collaborated slightly more with other females (statistically insignificant), the larger and broader difference seen was that in the years after 2015 collaboration with more females increased across all individuals. We examined these trends in our race data as well but due to the low number of total individuals represented there was no statistically significant differences (supplemental figure 1A).

### Female representation in neurosurgery is steadily increasing at the resident level, but board certifications still lag behind

From our dataset we utilized the American board of neurological surgery (ABNS) website to track year of board certification and compiled a rough timeline estimate of the growth of the neurosurgery field by decade (Figure 4A). Neurosurgery has had a steady growth rate over the last 40 years, but it wasn’t until the early 2000’s that female neurosurgeons from our dataset were getting consistently board certified (Figure 4A). Since then, the number of females board certified in neurosurgery has been steadily climbing (Figure 4B). To quantify the impact of a 2008 seminal white paper [26] on the field we pulled residency data [27,28] and plotted it along with board certification data (Figure 4B). Our data clearly shows a positive impact on the field in terms of female resident recruitment, however, even after two decades this has not translated in significant gains in female leadership in the field. Although many factors can influence this, to gain some insight on if longer academic progression time of male vs female neurosurgeons was one of the factors we tracked graduation to professor timelines of the top 15 performers. We plotted the mean time to promotion from graduation from residency to full professor for our top 15 male and female academic performers based on the available data and found an increase in the mean time to promotion in females (12.7 years in females to 9.3 years in males) but this change was not significant likely due to the very limited data (Sup Fig 2). To make these gains in the field sustainable in the long run more substantial faculty support and initiatives aimed at retention and development of promising female or URM faculty are needed (Figure 4C).

**Figure 4.**
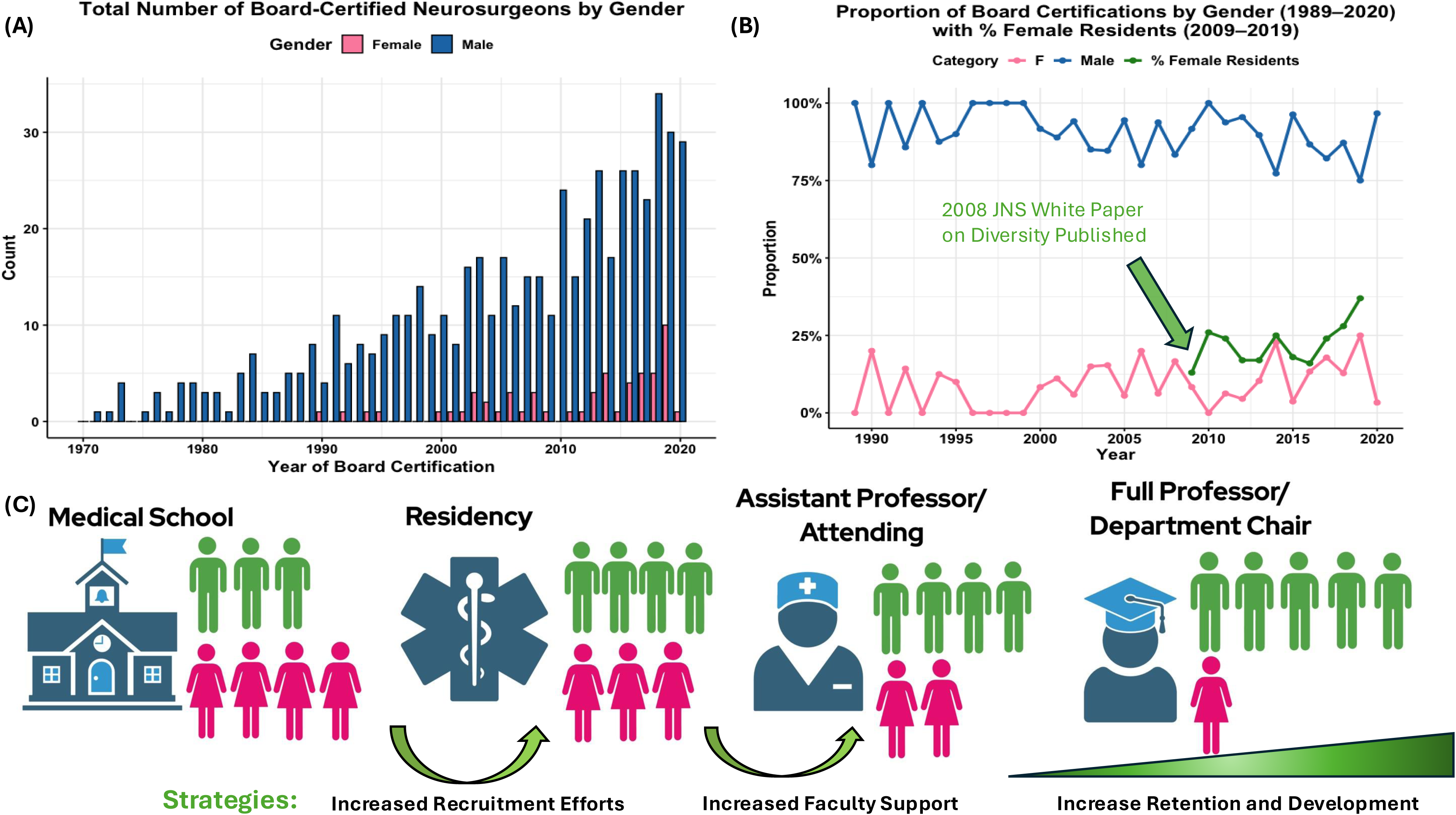
Gender trends in board certification of neurosurgeons over time: **A)** Bar plot of annual totals of board-certified neurosurgeons by gender per our web-scraped data. **B)** Line plot depicting yearly share of total certifications by gender and the percent of female neurosurgery residents (green). **C)** Cartoon depicting the loss of female faculty throughout the neurosurgery career pipeline along with strategies to combat this effect.

## Discussion

### Key Results

According to AAMC data in 2025, many entering medical school classes are routinely majority female, however, looking further up the ladder towards board certification and departmental leadership positions the effects have been less robust. Academic medicine, especially in surgical specialties, which plays a leading role in developing and implementing these policies, is still very white and male dominated. Our analysis demonstrates that although slow, these systemic policy changes and cultural shifts in medicine are having a positive impact in the profession of academic neurosurgery. In fact, over the last decade the field has trained more female neurosurgeons than the previous two decades combined. Although these numbers are positive there is still a wide gap in the field as more women and URM students than ever are now becoming physicians.

Our data suggests that this slower than expected change in the field of neurosurgery may be due to slower progression/growth of females and URM neurosurgeons in the academic sphere compared to non-URM male faculty. Although we definitively demonstrate more new female neurosurgeons are being trained than ever the currently established academically high ranking females are significantly less likely to receive a chair or department leadership position than males. In our dataset analyzing the correlation between leadership position and scholarly output we only identified 3/15 female chairs compared to 11/15 male chairs among the top 15 high preforming individuals. There was also no top performing male who was not at the level of full professor while we had 1 top performing female who had not yet attained this rank. We, and others [18–20], have found that even in 2025 there are a significant number of academic neurosurgery departments in the U.S that have no female faculty members at all. More female representation in leadership roles in academic neurosurgery would certainly drive more female collaborations potentially lifting the new and incoming larger proportion of females at the assistant professor levels. Furthermore, female leadership with real life experience of fundamental life events such as childbirth and caregiving will be more equipped to drive policies that can foster innovation in teaching, learning, and mentorship. This leadership will help to balance the demanding training schedule with sustainable family balance and make the field more sustainable for everyone in long run. Social networking phenomena such as homophily and unconscious bias also play a role in overall success of URM and female faculty. Research has shown that female physicians who enter surgical specialties have difficulties in establishing legitimacy, and face inequalities in career progression[9]. For example, a study analyzing surgical referral patterns found that male physicians were more likely to refer a patient to male surgeons compared to female surgeons, leading to lower patient volumes for female surgeons[29]. These types of ingrained human biases and perceptions can only be addressed by developing female and URM faculty and empowering them to rise to visible leadership positions.

The racial diversity of academic neurosurgery proved even harder to track and quantify than gender diversity since too few individuals were captured in our data to examine trends statistically. At all promotional levels we found URM physicians numbered in single digits. There is little comprehensive data describing the race of the neurosurgical workforce, however based on our data, along with others[14,21,25], we found that that URM physician matriculation into residency and transition to academic neurosurgery is likely stagnant or declining, especially when compared with corresponding increases in female representation.

### Limitations

We believe our survey data is representative of the broader academic neurosurgery workforce as our conclusions are similar to many other published reports[14,16,19,20,23,24]. However, due to the limitations of surveys we likely failed to capture some individuals, which in groups as small as women or URM individuals in academic neurosurgery, could have an impact on the data. We also had to rely on information provided by faculty webpages which are often outdated. There are also other nuanced leadership metrics such as program director, society presidents, society award recipients, research funding, journal editor in chief, etc. that could better inform leadership trends but are not addressed in this study to keep the results broad and generalizable. However other studies have clearly shown that all those more nuanced and advanced leadership metrics are very significantly Caucasian male dominant [30,31]. Another limitation of this study is that this data tries to assume trends but is largely based on single time point analyses when longitudinal study design would be more appropriate, however longitudinal examination would require repeated survey administration with significant study cost, although there have been studies attempting to do this on publicly available data which show similar conclusions to this work[32]. We are hopeful that this study provides an outline and incentive for national societies or ABNS to prospectively track the academic trajectory of graduating neurosurgeons in a longitudinal fashion as a “state of the field” type data collection.

## Conclusion

Progress, especially in academics, is slow and methodical. Our study definitively demonstrates gains in the field, but this gain has not yet reached leadership level. There has yet to be a year in the history of neurological surgery where a graduating residency class was less than 75% male [33] or where female leadership matched the female resident percentage. Addressing this discrepancy benefits everyone and having more diverse voices in leadership roles will only make neurosurgery stronger as a field.

## Data Availability

All data produced in the present study are available upon reasonable request to the authors

---

**Supplementary Figure 1:**
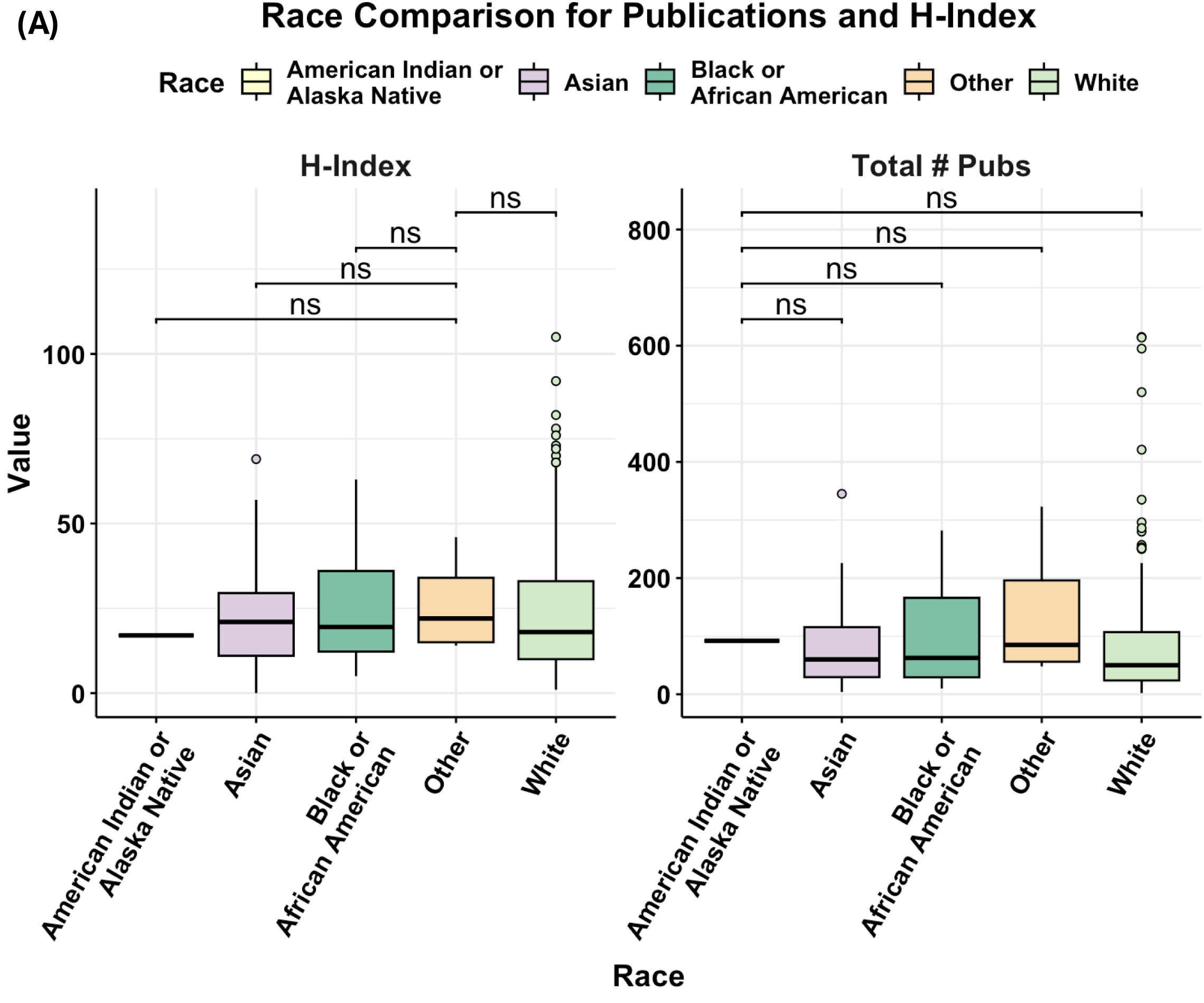
*Race-based comparison of academic productivity among neurosurgeons:* **A)** Boxplots comparing h-index and total publication count across racial groups.

**Supplementary Figure 2:**
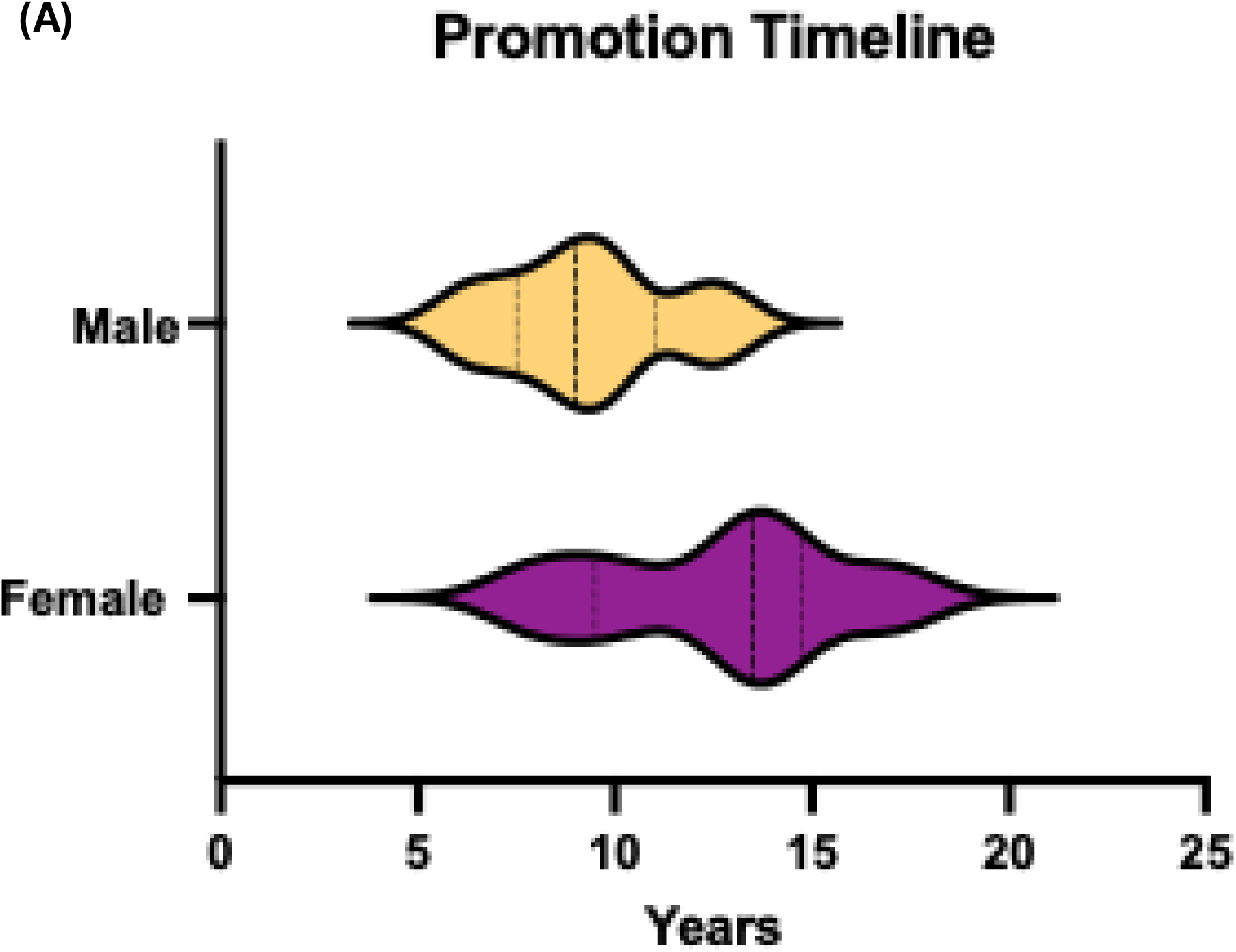
*Time to promotion from assistant to full professor across gender:* A) Time in years between assistant to full professor promotion expressed as violin plot from our top 15 male and female academic performers.

**Supplementary Table 1:** Questions asked on the survey distributed to US neurosurgical faculty.

