## Supplementary Figure 1 for "Expanding Faculty Representation in US Academic Neurological Surgery: Achievements and On-going Challenges"

| **Question** | **Response Type** |
| --- | --- |
| What is your preferred name, affiliated institution, and promotional rank? | Free response |
| What is your preferred gender? | Multiple Choice: A) Male, B) Female, C) Other (free response) |
| What is your race? | Multiple Choice: A) American Indian or Alaskan Native, B) Black or African American, C) Asian, D) White |
| What is your Ethnicity? | Multiple Choice: A) Hispanic Latino, B) Not Hispanic or Latino |
| Are you board Certified? | Multiple Chocie: A) Yes, B) No |
